# Registration, Publication, and Outcome Reporting among Pivotal Clinical Trials that Supported FDA Approval of High-Risk Medical Devices Before and After FDAAA

**DOI:** 10.1101/2021.02.28.21252619

**Authors:** Matthew J. Swanson, James L. Johnston, Joseph S. Ross

## Abstract

**Background:** Selective registration, publication, and outcome reporting of clinical trials distorts the primary clinical evidence that is available to patients and clinicians regarding the safety and efficacy of FDA-approved medical devices. The purpose of this study is to compare registration, publication, and outcome reporting among pivotal clinical trials that supported FDA approval of high-risk (Class III) medical devices before and after the U.S. Food and Drug Administration (FDA) Amendment Act (FDAAA) was enacted in 2007.

**Methods:** Using publicly available data from ClinicalTrials.gov, FDA summaries, and PubMed, we determined registration, publication, and reporting of findings for all pivotal clinical studies supporting FDA approval of new high-risk cardiovascular devices between 2005 and 2020, before and after FDAAA. For published studies, we compared both the primary efficacy outcome with the PMA primary efficacy outcome and the published interpretation of findings with the FDA reviewer’s interpretation (positive, equivocal, or negative).

**Results:** Between 2005 and 2020, the FDA approved 156 high-risk cardiovascular devices on the basis of 165 pivotal trials, 48 (29%) of which were categorized as pre-FDAAA and 117 (71%) as post-FDAAA. Post-FDAAA, pivotal clinical trials were more likely to be registered (115 of 117 (98%) vs 24 of 48 (50%); *p* < 0.001), to report results (98 of 115 (85%) vs 7 of 24 (29%); *p* < 0.001) on ClinicalTrials.gov, and to be published (100 or 117 (85%) vs 28 of 48 (58%); *p* < 0.001) in peer-reviewed literature when compared to pre-FDAAA. Among published trials, rates of concordant primary efficacy outcome reporting were not significantly different between pre-FDAAA trials and post-FDAAA trials (24 of 28 (86%) vs 96 of 100 (96%); *p* = 0.07), nor were rates of concordant trial interpretation (27 of 28 (96%) vs 93 of 100 (93%); *p* = 0.44).

**Conclusions:** FDAAA was associated with increased registration, results reporting, and publication for trials supporting FDA approval of high-risk medical devices. Among published trials, rates of accurate primary efficacy outcome reporting and trial interpretation were high and no different post-FDAAA.

## BACKGROUND

In 1976, Congress passed the Medical Device Amendments to the Food, Drug, and Cosmetic (FD&C) Act. This act established three classes for medical devices based on the regulatory controls necessary to provide reasonable assurance of their safety and efficacy.^1,2^ The most tightly regulated devices—those that support or sustain human life, are of substantial importance in preventing impairment of human health or could pose an unreasonable risk of illness or injury—are categorized as Class III (high-risk) devices. High-risk medical devices, such as stents, valves, sealants, and catheters, are regulated through the FDA’s Premarket Approval (PMA) pathway. Under the PMA pathway, device manufacturers are required to submit premarket clinical evidence that provides reasonable assurance of device safety and effectiveness.^3^ Pivotal clinical studies are generally the primary clinical evidence on which the FDA bases its approval decisions because they are designed to meet the aforementioned regulatory requirements, demonstrating both the safety and efficacy of the device for the intended use.^4^ Given the widespread use of high-risk medical devices in clinical practice, the quality and transparency of clinical data supporting their approval are of paramount importance to patient health and well-being.

Between January 2000 and December 2010, less than 50% of studies supporting PMA of novel, high-risk cardiovascular devices were published, and more than 30% of these publications presented primary endpoint results that were different, or could not be compared, to those in the corresponding FDA documents.^5^ These practices, known as selective publication and selective outcome reporting, distort the evidence available to patients and clinicians when making care decisions regarding the use of medical devices.^6,7^ In 2007, the U.S. FDA Amendment Act (FDAAA) was enacted, mandating clinical trial registration and results reporting on ClinicalTrials.gov for all ongoing and forthcoming trials of FDA-regulated products.^6^ It has been reported that post-enactment of FDAAA, pivotal efficacy trials supporting the approval of new drugs for cardiovascular disease, diabetes mellitus, and neuropsychiatric disease were significantly more likely to be registered, published, and have reported outcomes concordant with those submitted to FDA.^7,8^ However, no studies have examined the impact of FDAAA on the registration and reporting of clinical trials supporting FDA approval of medical devices.

Accordingly, we sought to characterize registration, results reporting, publication, and outcome reporting for pivotal studies supporting high-risk cardiovascular devices before and after the implementation of FDAAA. We focused on cardiovascular devices because they account for more than half of all FDA PMAs.^9^ Furthermore, we focused on pivotal studies because they are the definitive studies designed to evaluate medical device safety and effectiveness that are used as the basis of FDA’s regulatory decisions.^9,10^ The results of our study will inform future policy and regulatory efforts to ensure transparency and unbiased results reporting of the clinical trials supporting FDA approval of high-risk medical devices.

## METHODS

### Identification of high-risk cardiovascular medical devices

We identified novel, high-risk cardiovascular medical devices from the publicly accessible FDA PMA database (www.accessdata.fda.gov/scripts/cdrh/cfdocs/cfpma/pma.cfm) between January 1, 2005, and January 1, 2020, excluding automated external defibrillators (AEDs), studies that had missing data, and summaries that leveraged a meta-analysis for the pivotal study (Fig. 1).

**Figure 1.**
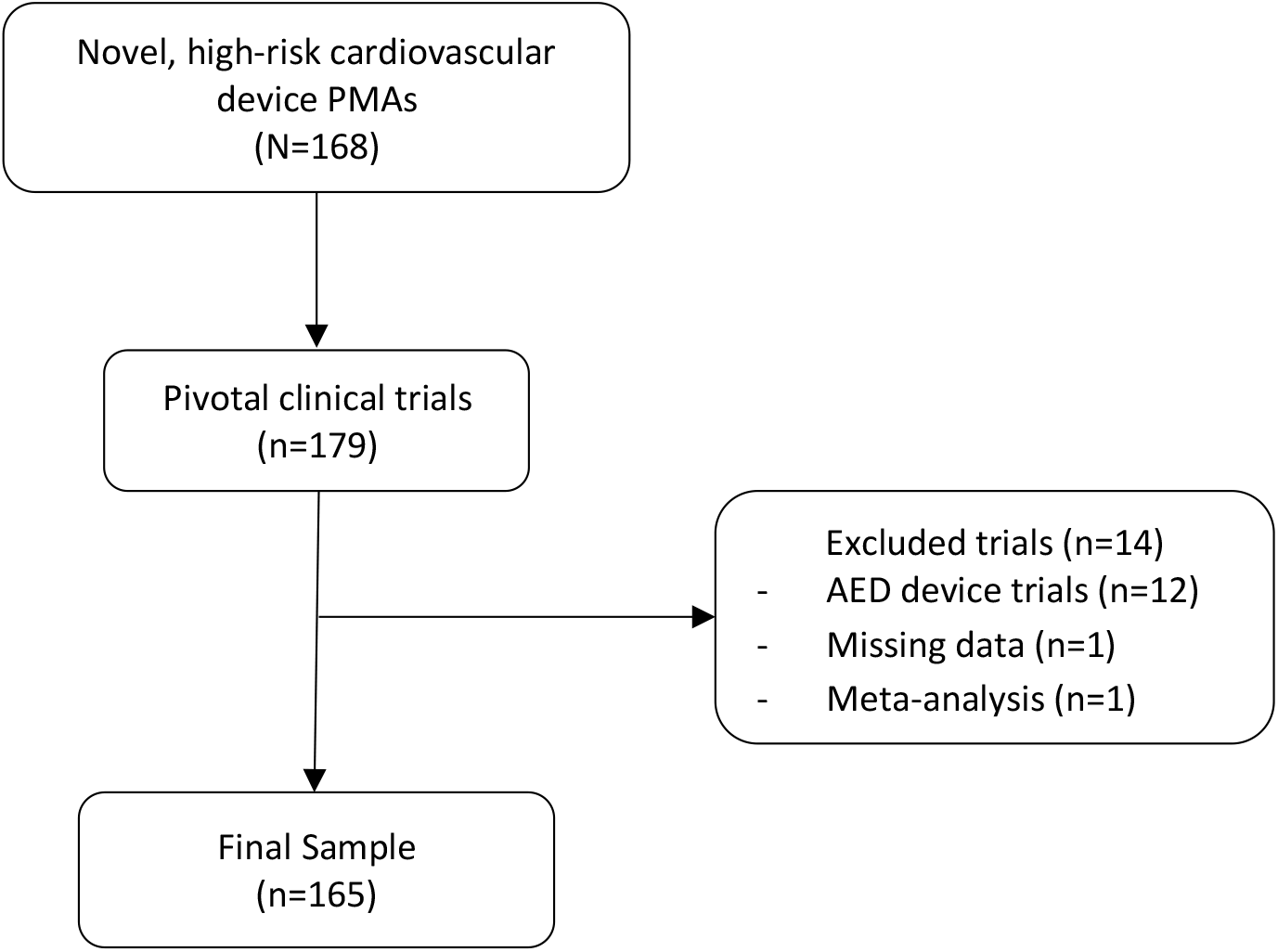
Sample construction of novel, high-risk cardiovascular devices approved by the US Food and Drug Administration between 2005 and 2020.

We excluded AEDs because FDA published a final order on January 29, 2015, stating that AED clinical study information can be leveraged from both published studies and clinical data previously submitted under the 510(k) process instead of requiring the conduct of a pivotal trial to support FDA approval.^11^ Otherwise, devices were selected if they met the following parameters: “Cardiovascular” under advisory committee and “Originals Only” under supplement type. All devices were characterized by the following using publicly available information on the FDA website: FDA review type (priority/standard), implantable designation (yes/no), life-sustaining designation (yes/no), and combination product (yes/no). Sponsor company management (public/private) was also determined by Google searching the sponsor company name along with “publicly traded,” “stock price,” “IPO,” or “privately held.”

### Characterization of pivotal clinical trials

For each device identified, we then identified the pivotal clinical studies that supported device approval from the “Summary of Safety and Effectiveness” documents. Pivotal clinical studies supporting approvals were categorized as pre-FDAAA if the clinical trial primary completion date was before December 26, 2007 (the date the policy took effect), in a manner described previously;^7^ all other studies were categorized as post-FDAAA. Also, we categorized pivotal trials by specific design characteristics: use of randomization (yes/no), use of blinded allocation (yes/no), primary efficacy endpoint (surrogate marker/clinical outcome or scale), and study center and patient enrollment numbers. Study characteristics and data were abstracted from the FDA summaries by one author (MJS); 10% of characterized trials were randomly selected and validated by a second author (JLJ). Any differences were reconciled by consensus.

### Identification of trials on ClinicalTrials.gov and published in the peer-reviewed literature

For each pivotal trial identified from FDA documents, we conducted a comprehensive search of ClinicalTrials.gov and PubMed’s listing of MEDLINE-indexed journals to identify any corresponding trial registration or publication, respectively. All document and website searches were performed during July 2020. Our search strategy included using and comparing clinical trial titles, product names, methods, number of study centers, enrollment numbers, primary efficacy endpoints, primary results, and study sponsors. While more recent FDA PMAs include ClinicalTrials.gov registration hyperlinks and ClinicalTrials.gov manually and automatically indexes corresponding publications of results to their registration by National Clinical Trial (NCT) number, these identification numbers did not reliably identify pivotal trial registrations and publications for older PMAs, consistent with prior reviews.^12^ From among publications identified in PubMed, abstracts and conference reports were excluded. Publications reporting multiple studies, such as reviews and meta-analyses, were also excluded unless the results of each study were analyzed and discussed individually at the level of detail as one would expect from a full-length publication.

### Comparison to corresponding publications

First, for each pivotal trial for which a publication was identified, we compared the primary effectiveness endpoint specified in the FDA documents with the effectiveness endpoint specified as primary in the publication. If there was more than one primary effectiveness endpoint reported in the FDA documents, we verified the one that matched the primary endpoint specified in the publication. If none of the specified primary endpoints matched, we categorized the primary effectiveness outcomes reported as discordant. If one matched, we determined whether the primary effectiveness endpoint result reported in the FDA documents was the same as the result reported in the publication. The outcomes reported were categorized as concordant if they were an exact numerical match or if there was a relative difference of less than 5% to the FDA PMA, and discordant if they were not. Second, for each pivotal trial for which a publication in the peer-reviewed literature was identified, we compared the overall study interpretation between the two sources. The overall interpretation was categorized as positive, equivocal, or negative based on the FDA officer’s language in the “Effectiveness Conclusions” and “Overall Conclusions” subsections of the “Summary of Safety and Effectiveness” document and the author’s language in the conclusion of the publication; the FDA and publication interpretation were categorized as concordant or discordant.

### Statistical analysis

We determined the rate of ClinicalTrials.gov registration, ClinicalTrials.gov results reporting, and PubMed publication for all identified pivotal trials, overall and stratified by device and design characteristics. We then determined the overall rate of concordant primary outcome reporting between the FDA PMA summaries and corresponding publications, as well as the overall rate of concordance between the FDA PMA reviewer’s interpretations and the trial publication’s interpretations. Summary statistics were calculated for each comparison, presented as numbers, percentages, means, standard deviations, and ranges, as appropriate. Chi-square and two-tailed Fisher exact tests were used to compare rates pre- and post-FDAAA of registration, results reporting, publication, concordant outcome reporting, and concordant interpretation, as appropriate. All statistical tests were two-tailed and used the Bonferroni method to adjust P values to account for the five comparisons; statistical significance was set at P ≤ 0.01. Analyses were performed using Microsoft Excel (version 16.35) and SPSS (version 27).

This study was prepared in accordance with the Strengthening the Reporting of Observational Studies in Epidemiology (STROBE) reporting guideline for cross-sectional studies.^13^ The study did not require institutional review board approval or patient informed consent because it was based on publicly available information and involved no patient records.

## RESULTS

Between 2005 and 2020, the FDA approved 156 novel, high-risk cardiovascular devices (Table 1). Among these, 16 (10%) approvals were designated for priority review, 84 (54%) were life-sustaining, 123 (79%) were implantable, 29 (19%) were combination products, and 60 (38%) had private sponsor company management.

**Table 1.** Novel, high-risk cardiovascular devices approved by the US FDA between 2005 and 2020.

| Novel approvals (N = 156) | Number (%) |
| --- | --- |
| FDA review pathway<sup>a</sup> |  |
| Priority | 16 (10) |
| Standard | 140 (90) |
| Life-sustaining designation<sup>a</sup> |  |
| Life-sustaining | 84 (54) |
| Non-life-sustaining | 72 (46) |
| Implantable designation<sup>a</sup> |  |
| Implantable | 123 (79) |
| Non-implantable | 33 (21) |
| Combination Product<sup>a</sup> |  |
| No | 127 (81) |
| Yes | 29 (19) |
| Company Management |  |
| Public | 96 (62) |
| Private | 60 (38) |

*FDA* US Food and Drug Administration
<sup>a</sup>FDA designation

We identified a total of 179 pivotal clinical trials supporting these 156 approvals, of which 165 met our inclusion criteria (Fig. 1), among which 48 (29%) were categorized as pre-FDAAA and 117 (71%) as post-FDAAA. Overall, 59 (36%) of these pivotal trials were randomized, 21 (13%) were blinded, 103 (62%) were from publicly-held companies, 19 (12%) supported devices with priority review status, 32 (19%) supported combination products, 132 (80%) supported implantable devices, and 89 (54%) supported life-sustaining devices.

### Trial registration, results reporting, and publication

Among the 165 pivotal trials that met our inclusion criteria, 139 (84%) were registered on ClinicalTrials.gov, 105 (76%) had results posted on ClinicalTrials.gov, and 128 (78%) were published in the peer-reviewed literature. Compared to pre-FDAAA pivotal trials, post-FDAAA trials were more likely to be registered on ClinicalTrials.gov (115 of 117 (98%) vs 24 of 48 (50%); *p* < 0.001), to report results on ClinicalTrials.gov (98 of 115 (85%) vs 7 of 24 (29%); *p* < 0.001), and to be published (100 of 117 (85%) vs 28 of 48 (58%); *p* < 0.001) (Table 2). Trials registered on ClinicalTrials.gov were more likely to be published than those not registered (121 of 139 (87%) vs 7 of 26 (27%); *p* < 0.001). Pre-FDAAA, implantable designation was associated with greater likelihood of registration (21 of 36 (58%) vs 3 of 12 (25%); p = 0.05) and life-sustaining designation was associated with greater likelihood of results reporting or publication (18 of 24 (75%) vs 11 of 24 (46%); p = 0.04; Table 3). Post-FDAAA, there were no significant differences in registration, results reporting, or publication when stratified by device or trial design characteristics (Table 4).

**Table 2.**
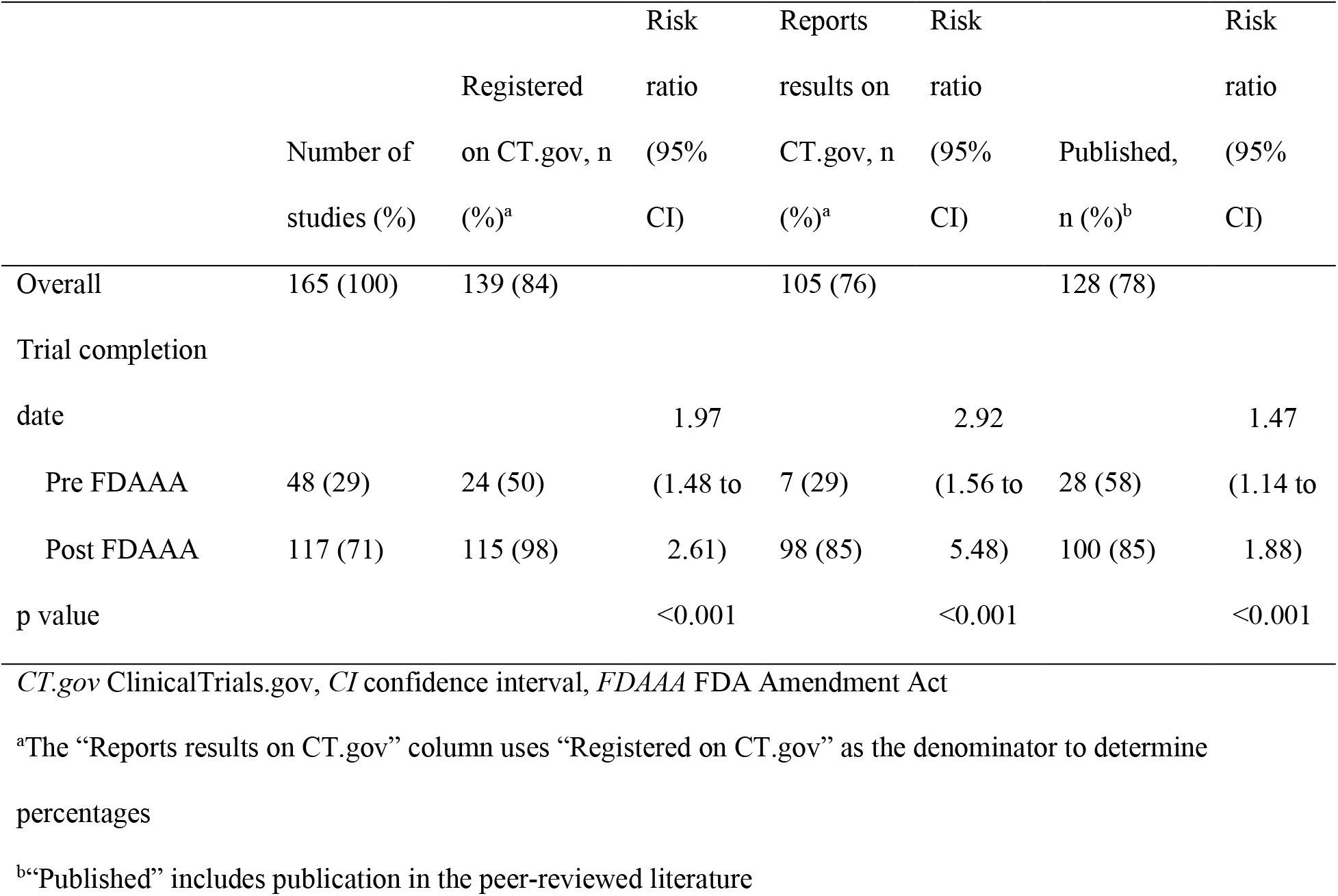
Registration, results reporting, and publication of clinical trials supporting FDA cardiovascular device approvals between 2005 and 2020, pre- and post-FDAAA (n=165).

**Table 3.**
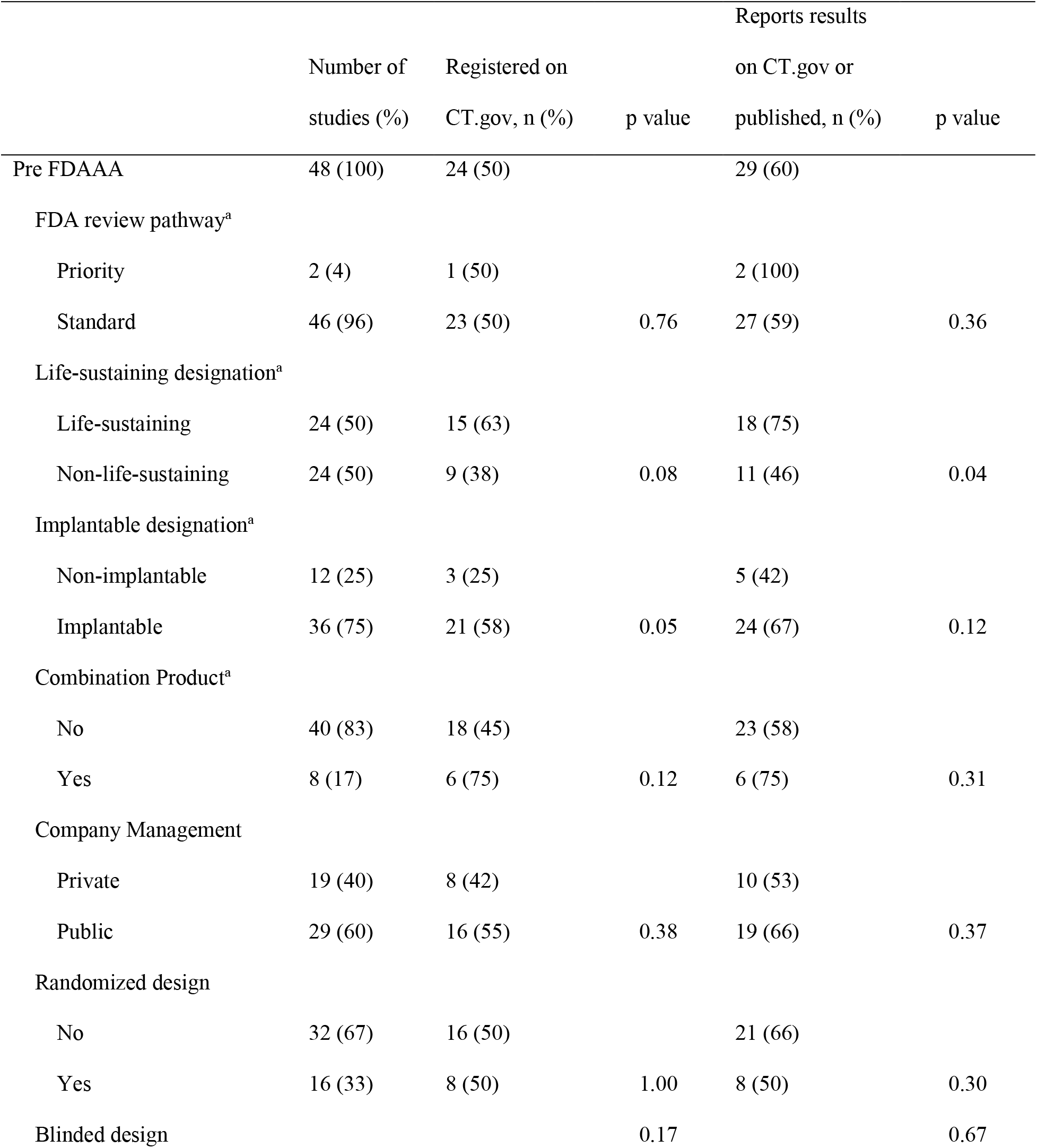
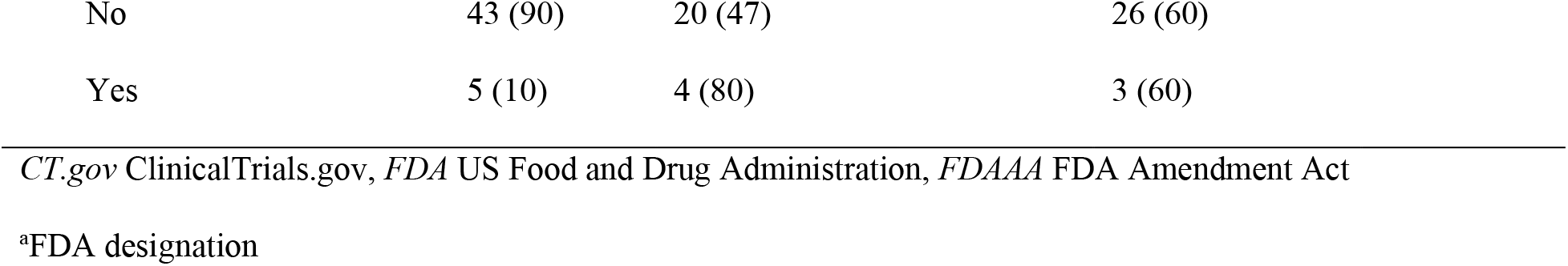
Registration and results reporting or publication of clinical trials supporting FDA cardiovascular device approvals between 2005 and 2020, stratified by study and device characteristics, pre-FDAAA (n=48).

**Table 4.**
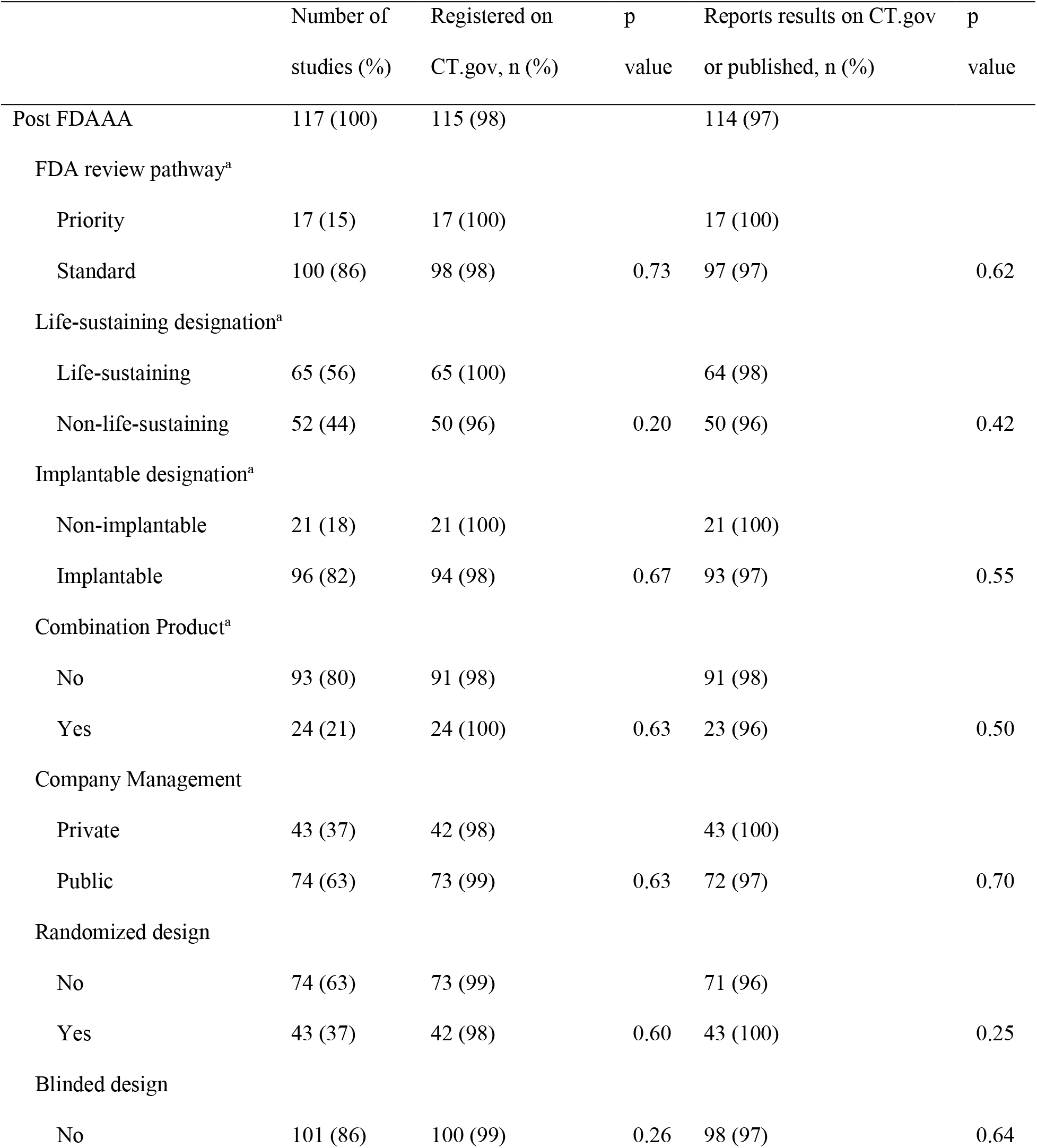
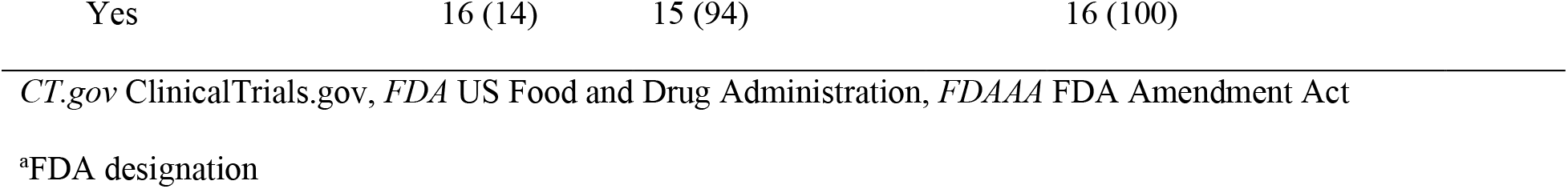
Registration and results reporting or publication of clinical trials supporting FDA cardiovascular device approvals between 2005 and 2020, stratified by study and device characteristics, post-FDAAA (n=117).

### Concordant outcome reporting and trial interpretation

A primary effectiveness outcome and main result were reported in the FDA documents for all pivotal trials, pre- and post-FDAAA. Overall, 37 of 165 (22%) trials were not published, precluding primary effectiveness outcome comparison. Among 120 of 128 (94%) published trials, the primary endpoint specified in the FDA documents was the same endpoint specified as primary in the publication; among these, the primary endpoint result reported in the FDA documents was the same or within 5% of the result reported in the publication for 120 (100%) trials. Rates of concordant primary effectiveness outcome reporting were not significantly different between published pre-FDAAA and post-FDAAA trials (24 of 28 (86%) vs 96 of 100 (96%); *p* = 0.07) (Table 5).

**Table 5.** Outcome reporting and interpretation concordance of published clinical trials supporting FDA cardiovascular device approvals between 2005 and 2020, pre- and post-FDAAA (n=128).

|  | Number of<br>published studies<br>(%) | Concordant outcome<br>reporting, n (%) | Risk ratio<br>(95% CI) | Concordant<br>interpretation, n (%) | Risk ratio<br>(95% CI) |
| --- | --- | --- | --- | --- | --- |
| Overall | 128 (100) | 120 (94) |  | 120 (94) |  |
| Trial completion date |  |  |  |  |  |
| Pre FDAAA | 28 (22) | 24 (86) | 1.12 (0.96 | 27 (96) | 0.96 (0.88 |
| Post FDAAA | 100 (78) | 96 (96) | to 1.31) | 93 (93) | to 1.06) |
| p value |  |  | 0.07 |  | 0.44 |
| *CI* confidence interval, *FDAAA* FDA Amendment Act |  |  |  |  |  |

Among the 48 pre-FDAAA trials, FDA reviewers characterized 45 (94%) as positive, 2 (4%) as equivocal, and 1 (2%) as negative, whereas among the 117 post-FDAAA trials, FDA reviewers characterized 105 (90%) as positive, 3 (3%) as equivocal, and 9 (8%) as negative (Fig. 2). Overall, 116 of 150 (77%) positive trials were published, 4 of 5 (80%) equivocal trials, and 8 of 10 (80%) negative trials. Among published trials, rates of concordant interpretation were not significantly different between pre-FDAAA trials and post-FDAAA trials (27 of 28 (96%) vs 93 of 100 (93%); *p* = 0.44). All discordant trials were published in a manner that conveyed a more positive interpretation than that of the FDA reviewer. No trials were published in a manner that conveyed a more negative interpretation than that of the FDA reviewer.

**Figure 2.**
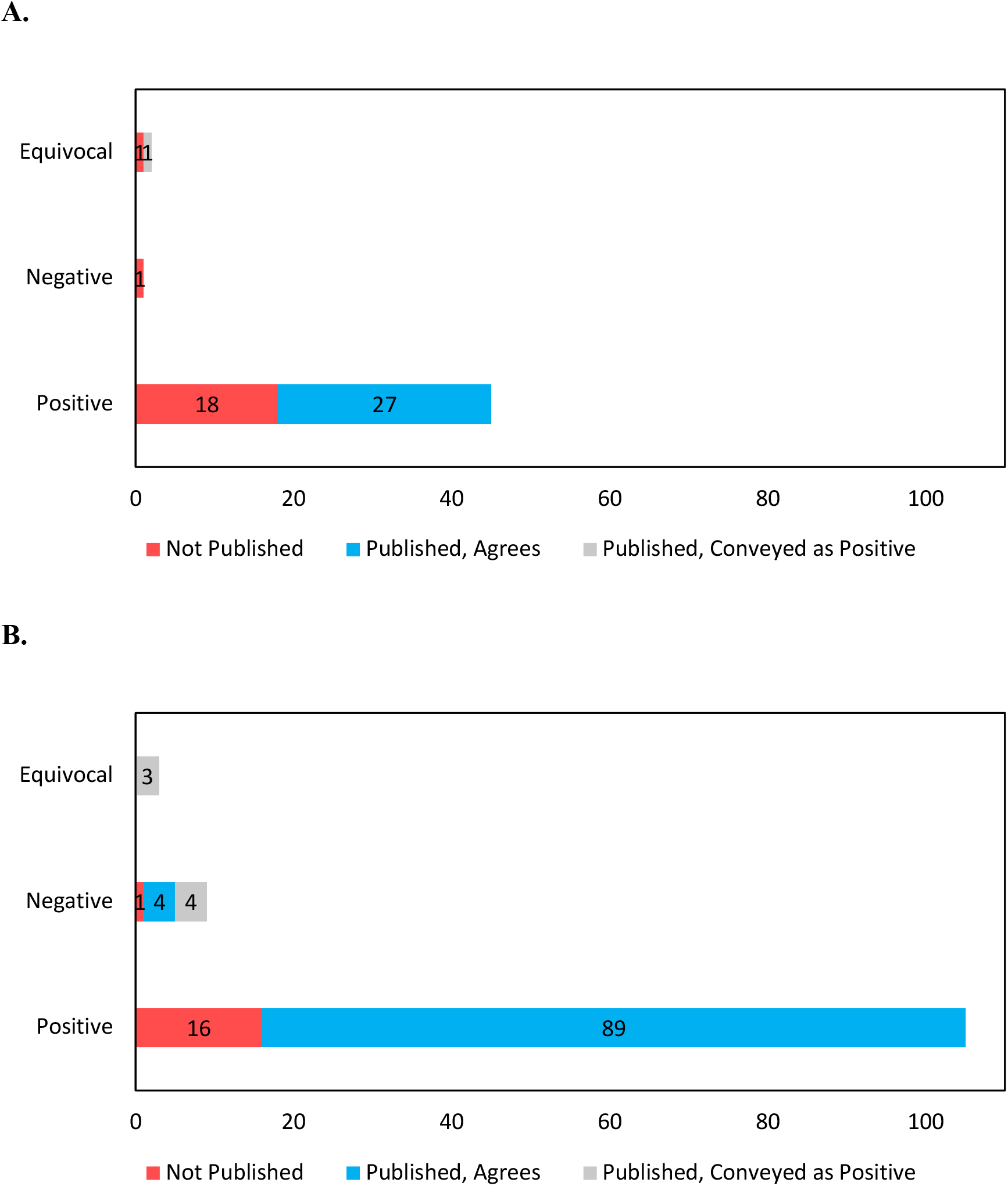
FDA reviewer trial interpretation and publication, along with published interpretation of the trial findings, for novel cardiovascular devices approved by the US FDA between 2005 and 2020, pre- and post-FDAAA. FDA reviewer trial interpretation as positive, equivocal, or negative. (A) Pre-FDAAA; (B) Post-FDAAA.

## DISCUSSION

In our study of all pivotal studies supporting high-risk cardiovascular devices approved by the FDA through the PMA pathway from 2005 to 2020, we found that implementation of FDAAA, which mandated clinical trial registration and results reporting, was associated with higher rates of pivotal trial registration, results reporting, and publication, but no differences in the accuracy of outcome reporting and trial interpretation among published studies. These results suggest that the legislation has improved transparency and unbiased results reporting of clinical trials, potentially mitigating selective publication and outcome reporting, which can thereby ensure that patient care decisions are based on more complete and accurate research.

Our study demonstrates that 98% of post-FDAAA trials were registered on ClinicalTrials.gov, 85% reported their results on ClinicalTrials.gov, and 85% were published in the peer-reviewed literature. These rates are high but lag behind those reported for new drugs.^7,8^ Impressive clinical trial registration and results reporting after FDAAA enactment was expected given the explicit requirement to require trial registration among all trials investigating FDA-regulated products.

However, there is still room for improvement because 2% of post-FDAAA trials remain unregistered on ClinicalTrials.gov, 15% have not posted their results on ClinicalTrials.gov, and 15% remain unpublished in the peer-reviewed literature. As required by law, all of these rates should be 100%. Follow-up studies will be needed to see whether these rates persist or improve.

Among post-FDAAA trials, publication rate in the peer-reviewed literature was higher when compared to the rate of 80% observed among studies supporting FDA approval of novel, high-risk cardiovascular devices between January 2011 and December 2013.^14^ Although it continues to greatly exceed the rate of 49% observed for trials supporting FDA-approved, high-risk cardiovascular devices between January 2000 and December 2010,^5^ given that these pivotal trials represent the best evidence of medical device safety and effectiveness, there is no reason that the clinical and research community should not expect a publication rate of 100%. Also, 96% of published studies reported primary effectiveness outcomes in a manner concordant with FDA reviews, which is nearly identical to a prior study of medical device publication and results reporting.^14^ Our rate may be more representative of concordant results reporting post-FDAAA because we analyzed 15 years of FDA approvals post-FDAAA while the aforementioned study analyzed three years post-FDAAA. Nonetheless, our results showed higher rates than the initial study of selective reporting for medical devices that reported a rate of 69% for both identical and similar primary endpoint reporting, which when binned most closely matched our “concordant outcome reporting” statistic, in pivotal high-risk cardiovascular device trials between 2000 and 2010.^5^

We found that 93% of published trial interpretations were concordant with FDA reviews, which is slightly lower than a prior study of medical device publication and results reporting^14^ but again may be more representative of concordant interpretation reporting post-FDAAA because of our larger sample size. There were no significant differences in rates of publication when examining device and design characteristics among pre- and post-FDAAA trials, which differs from a previous report of cardiovascular device research that found both publicly-held held company sponsors and life-sustaining device designation to be associated with the likelihood of publication.^14^

There are several limitations to be considered in the interpretation of our findings. First, we only looked at trials supporting FDA approval of high-risk cardiovascular devices. Our results may not be generalizable to all high-risk medical devices and should be confirmed for FDA approvals in other therapeutic areas. Second, our study is cross-sectional and observational, we can only establish associations, not causal conclusions, about the impact of FDAAA. Third, we limited our search of trial registration to ClinicalTrials.gov and excluded the use of other clinical trial registration sites. That said, U.S. law requires trial registration of FDA-regulated products on ClinicalTrials.gov. Finally, our study was focused on the reporting and publication of primary efficacy endpoints and interpretations and did not examine reporting or publication of secondary efficacy and safety endpoints.

## CONCLUSION

FDAAA was associated with higher rates of clinical trial registration on ClinicalTrials.gov, results reporting, and publication in the peer-reviewed literature for trials supporting FDA approval of high-risk cardiovascular medical devices. Among published trials, rates of accurate primary efficacy outcome reporting and trial interpretation were high and no different post-FDAAA. These findings have important implications for understanding the potential impact of the FDAAA and informing future policy and regulatory efforts to ensure transparency and unbiased results reporting of the clinical trials supporting FDA approval of high-risk medical devices.

## Supporting information

STROBE Checklist

## Data Availability

The datasets used and/or analyzed during the current study are available from the corresponding author on reasonable request.

## LIST OF ABBREVIATIONS

FDA: Food and Drug Administration
FDAAA: Food and Drug Administration Amendment Act
PMA: Premarket Approval
AED: Automated External Defibrillator
NCT: National Clinical Trial
STROBE: Strengthening the Reporting of Observational Studies in Epidemiology

## DECLARATIONS

### ETHICS APPROVAL AND CONSENT TO PARTICIPATE

Because our examination of trial publications did not involve human subjects, ethics committee review was not required by the Yale University Human Research Protection Program. Consent to participate not applicable.

## CONSENT FOR PUBLICATION

Not applicable.

## AUTHORS’ CONTRIBUTIONS

MJS was responsible for the conception, design, acquisition of data, analysis and interpretation of the data, and drafting and revision of the manuscript. JLJ was responsible for the acquisition of data, analysis and interpretation of the data, and critical revisions for important intellectual content. JSR was responsible for the conception, design, analysis, and interpretation of the data, and drafting and revision of the manuscript. JSR provided supervision. All authors read and approved the final manuscript.

## ACKNOWLEDGEMENTS

Not applicable.

## REFERENCES

1. Medical device amendments of 1976. <https://www.govinfo.gov/content/pkg/STATUTE->90/pdf/STATUTE-90-Pg539.pdf.

2. Federal food, drug, and cosmetic act (FD&C act). https://catalog.archives.gov/id/299847.

3. Premarket approval (PMA). <https://www.fda.gov/medical-devices/premarket->submissions/premarket-approval-pma.

4. Design considerations for pivotal clinical investigations for medical devices. <https://www.fda.gov/media/87363/download.>

5. Chang L, Dhruva SS, Chu J, Bero LA, Redberg RF. Selective reporting in trials of high risk cardiovascular devices: Cross sectional comparison between premarket approval summaries and published reports. BMJ. 2015;350:h2613. doi: 10.1136/bmj.h2613 [doi].

6. The food and drug administration amendments act of 2007. Public law 110-85. <https://www.gpo.gov/fdsys/pkg/PLAW-110publ85/pdf/PLAW-110publ85.pdf.>

7. Phillips AT, Desai NR, Krumholz HM, Zou CX, Miller JE, Ross JS. Association of the FDA amendment act with trial registration, publication, and outcome reporting. Trials. 2017;18(1):333–3. doi: 10.1186/s13063-017-2068-3 [doi].

8. Zou CX, Becker JE, Phillips AT, et al. Registration, results reporting, and publication bias of clinical trials supporting FDA approval of neuropsychiatric drugs before and after FDAAA: A retrospective cohort study. Trials. 2018;19(1):581. https://doi.org/10.1186/s13063-018-2957-0. xdoi: 10.1186/s13063-018-2957-0.

9. Rathi VK, Krumholz HM, Masoudi FA, Ross JS. Characteristics of clinical studies conducted over the total product life cycle of high-risk therapeutic medical devices receiving FDA premarket approval in 2010 and 2011. JAMA. 2015;314(6):604–612. doi: 10.1001/jama.2015.8761 [doi].

10. Downing NS, Aminawung JA, Shah ND, Krumholz HM, Ross JS. Clinical trial evidence supporting FDA approval of novel therapeutic agents, 2005-2012. JAMA. 2014;311(4):368–377. doi: 10.1001/jama.2013.282034 [doi].

11. Effective date of requirement for premarket approval for automated external defibrillator systems. <https://www.federalregister.gov/documents/2015/01/29/2015-01619/effective-date-of->requirement-for-premarket-approval-for-automated-external-defibrillator-systems.

12. Bashir R, Bourgeois FT, Dunn AG. A systematic review of the processes used to link clinical trial registrations to their published results. Syst Rev. 2017;6(1):123–3. doi: 10.1186/s13643-017-0518-3 [doi].

13. von Elm E, Altman DG, Egger M, Pocock SJ, Gotzsche PC, Vandenbroucke JP. The strengthening the reporting of observational studies in epidemiology (STROBE) statement: Guidelines for reporting observational studies. <https://www.equator-network.org/reporting->guidelines/strobe/.

14. Phillips AT, Rathi VK, Ross JS. Publication of clinical studies supporting FDA premarket approval for high-risk cardiovascular devices between 2011 and 2013: A cross-sectional study. JAMA Intern Med. 2016;176(4):551–552. doi: 10.1001/jamainternmed.2015.8590 [doi].

